# Predicting Trends of Coronavirus Disease (COVID-19) Using SIRD and Gaussian-SIRD Models

**DOI:** 10.1101/2020.11.29.20240499

**Authors:** Ahmad Sedaghat, Amir Mosavi

## Abstract

Eruption of COVID-19 patients in 215 countries worldwide have urged for robust predictive methods that can detect as early as possible size and duration of the contagious disease and also providing precision predictions. In many recent literatures reported on COVID-19, one or more essential parts of such investigation were missed. One of crucial elements for any predictive method is that such methods should fit simultaneously as many data as possible; these data could be total infected cases, daily hospitalized cases, cumulative recovered cases and deceased cases and so on. Other crucial elements include sensitivity and precision of such predictive methods on amount of data as the contagious disease evolved day by day. To show importance of these aspects, we have evaluated the standard SIRD model and a newly introduced Gaussian-SIRD model on development of COVID-19 in Kuwait. It is observed that SIRD model quickly pick up main trends of COVID-19 development; but Gaussian-SIRD model provides precise prediction at longer period of time.

## I. Introduction

COVID-19 outbreak was first reported as contiguous disease created by novel corona virus in Wuhan, China in late December 2019. Soon after, the world has faced with the most rapidly transmitting pandemic disease which halted our normal way of living with 11,312,265 infected and 531,257 deceased cases worldwide on 5th July 2020 [1].

Quick and precise prediction of endemic/pandemic contagious diseases dynamics is very important for health authorities and governments to tackle the disease in the most efficient and economical way. COVID-19 have caused many businesses broken down and several million people worldwide out of their jobs. SIR model is well known and widely used method introduced by Kermack andMcKendrick [2] in epidemiological studies. SIR method is based on 3-set of ordinary differential equations expressing susceptible, infected, and removed populations in a community with constant total population. SIR model assumes that sex, age, social behaviour, and similar factors has no effects on development of a contagious disease. Exact solutions to SIR model may improve predictive capabilities of this method by employing optimization techniques. However, there are a limited exact solution to SIR model in simplified forms. Bailey [3] provided a simplified form of SIR equations. Bohner et al. [4] reported exact solution to Bailey’s model of SIR equations. In his work, recovered population is not considered in susceptible and infection equations. Harko et al. [5] considered birth and death rates in an exact solution to SIR model; however, the validity of their method was not evaluated against actual epidemic data. Maliki [6] and Shabbir et al. [7] reported separately exact solutions to some simplified SIS and SIR equations. These simplified models do not include the removed populations and only consider susceptible and infected populations.

In the present work, SIRD model consist of susceptible, infected, recovered, and deceased populations correspond to 4-set of ordinary differential equations (ODE) are considered. In the new Gaussian-SIRD model, a Gaussian function is assumed for infected population; hence, 4-set of explicit analytical solutions are found for SIRD populations. MATLAB optimization are applied to fit the SIRD and Gaussian-SIRD models with COVID-19 data in Kuwait. Goodness of fit functions for both methods were evaluated using the coefficient of determination (R2). Sensitivity and precision of both methods are compared for COVID-19 development in Kuwait and advantageous and pitfalls of each method are discussed and conclusions of this study are drawn.

## II. Materials and methdos

To tackle a pandemic, correct evaluation of transmission and growth rates of disease and affected populations are essential. SIR model is the widely used method in epidemiology studies. In SIR type models, total population size is considered constant during an endemic/pandemic. SIR model does not consider human factors including sex of patients, age and social behaviour of patients, and also location of infected cases. These are shortcomings of SIR model for predicting pandemics such as COVID-19 for which strong dependency observed in individual age or sex and social behaviour [8, 9].

### A. SIRD Model

A modified version of SIR model to include deceased population referred as SIRD model here. SIRD model consist of 4-set of ordinary differential equations on susceptible, infected, recovered, and deceased population as follows [10, 11]:

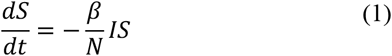

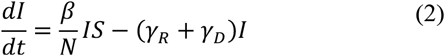

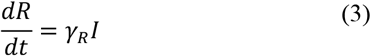

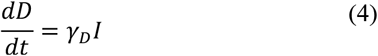

It is assumed that initial total population (*N*) is constant during an endemic/pandemic:

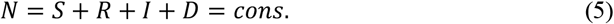

In equations (1-5), *β* is the transmission rate, *γ*_*R*_ is the recovery rate, *γ*_*D*_ is the death rate in an endemic/pandemic. *γ* (= *γ*_*R*_ + *γ*_*D*_) expresses the removing rate of recovered and deceased populations from susceptible population. An important factor in studying epidemiological field is called the reproduction number *R*_0_ (= *β*/*γ*), which indicates severity of an outbreak. If *R*_0_ value is larger than one then it is expected that infection will be spread in susceptible population. The larger *R*_0_ will indicate harder to control of spread of infection.

Observing equations (1-5), an analytical solution to equation (2) for infected population will be sufficient to determine explicit solutions for all other populations in equations (1-5). In the next section, we introduce an analytical solution to SIRD model assuming a Gaussian distribution for infected populations.

### B. Gaussian-SIRD Model

Normal or Gaussian distribution function is widely used in statistics to deal with continuous probability of real value of a random variable [12]. The normal probability density function is often used in studying natural and social subjects when real value of the random variables is not known. Gaussian distribution function of two parameter family (*μ, σ*) may be reformulated for infected population of a pandemic as follows [13]:

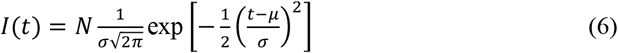

In equation (6), *I*(*t*) is the Gaussian normal distribution function of daily infected population, *N* is the total population, *μ* is the mean (or expectation) of the distribution and *σ* is the standard deviation. The *μ*, and *σ* are model parameters and can be obtained by fitting Gaussian equation (6) with infectious population from a pandemic data.

Exact solutions to recovered (R) and deceased (D) is simply obtained by plugging equation (6) into equations (3) and (4) to obtain cumulative solution as follows [13]:

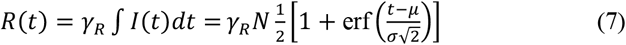

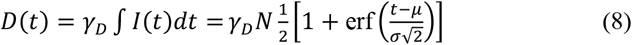

In equations (6) and (7), the error function erf(*x*) is a special function and is evaluated numerically.

Substituting equation (6) into equation (1), one may obtain a solution for susceptible population as follows:

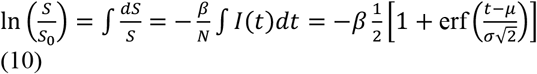

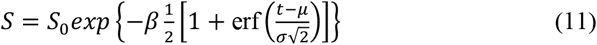

By changing the total susceptible population (N) in SIRD model, the recovered and deceased will be massively changed. To fine-tune expected recovered and deceased cases, we propose to use variable recovery rate (*γ*_*R*_) and deceased rate (*γ*_*D*_) as follows:

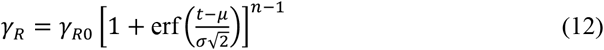

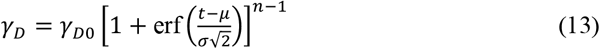

The power factor (n) is any number and can be adequately fine-tuned to best fit the recovered and deceased populations data.

The above set of analytical equations (6-8, 11) provide a simple analytical solution to an endemic/pandemic data. We have applied optimization to get best value of coefficients in SIRD and Gaussian-SIRD models. MATLAB algorithm (lsqcurvefit) [14] was applied to find best coefficients in SIRD model equations (1-5). Also, MATLAB algorithm (fminsearch) [15] was used to find best fit coefficients in Gaussian-SIRD model equations (6-8, 11). The goodness of fitted COVID-19 data are examined using the coefficient of determination (R^2^) [16].

### C. Evaluation on goodness of fit: Regression coefficient

Regression coefficient (R^2^) is a statistical measure to compare predicted values (*y*) from SIRD or Gaussian-SIRD models here against actual data (*x*) for COVID-19. The evaluation of R^2^ is done separately for each population: susceptible, infected, recovered, and death as follows [16]:

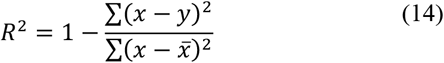

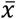 in equation (14) is the average of actual COVID-19 data values. Bette fit functions indicated the regression coefficient (R^2^) close to unity.

### D. Sensitivity analysis

To measure sensitivity of a predictive method, a comparison of actual data values (*x*) and predicted values (*y*) can be expressed in percentage of error as follows [16]:

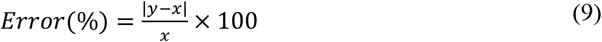

Error percentage values closer to zero provides a guide on sensitivity and precisions of these methods.

## III. Results

We have investigated sensitivity of SIRD and Gaussian-SIRD models on predicting population size and peak day of infectious using 20, 40, 60, 80, 100, and 116 subsequent days of COVID-19 data in Kuwait [17]. Results of optimized SIRD [18] and Gaussian-SIRD models are presented and compared in Fig. 1 for susceptible, infected, recovered, and deceased populations.

**Figure 1.**
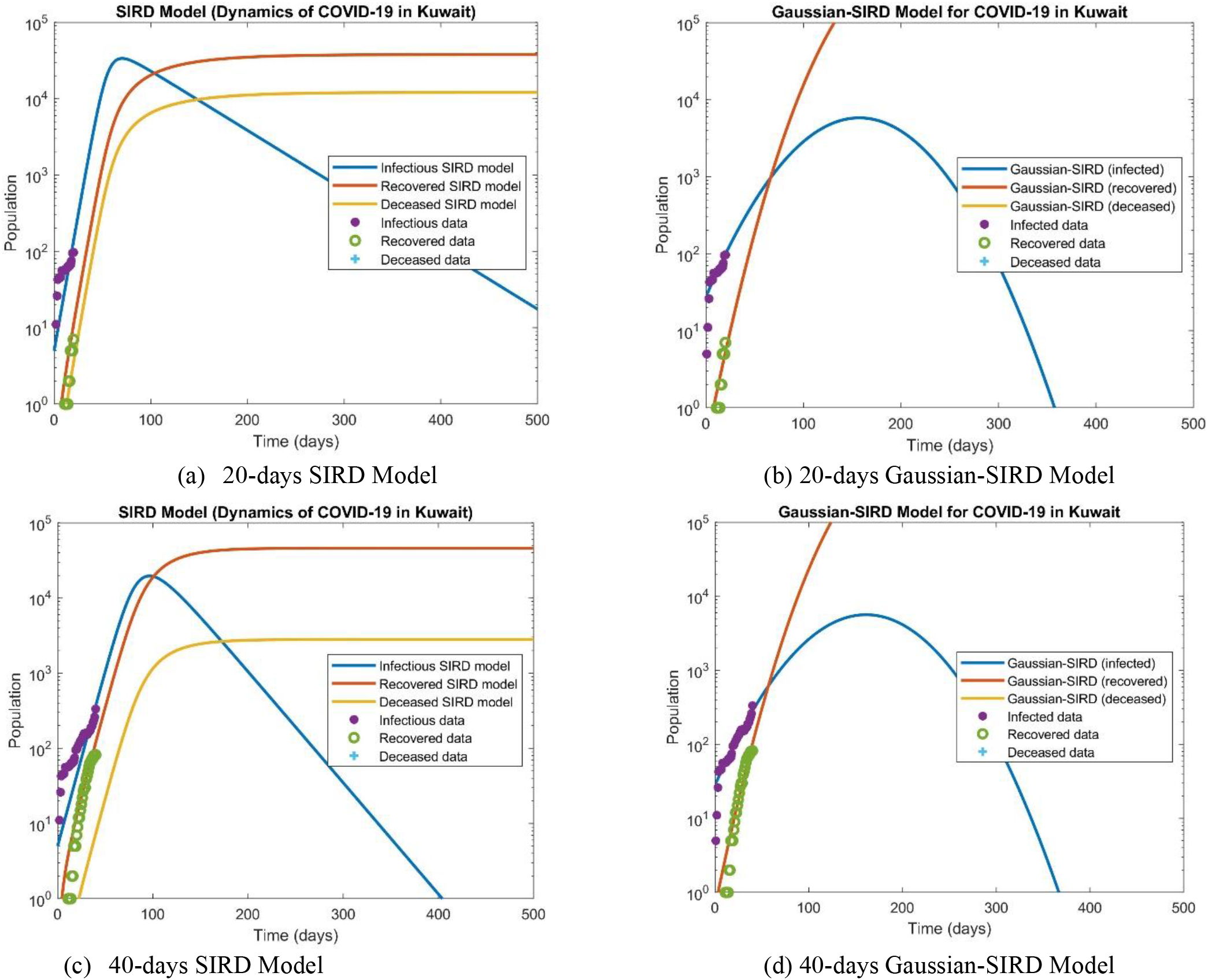
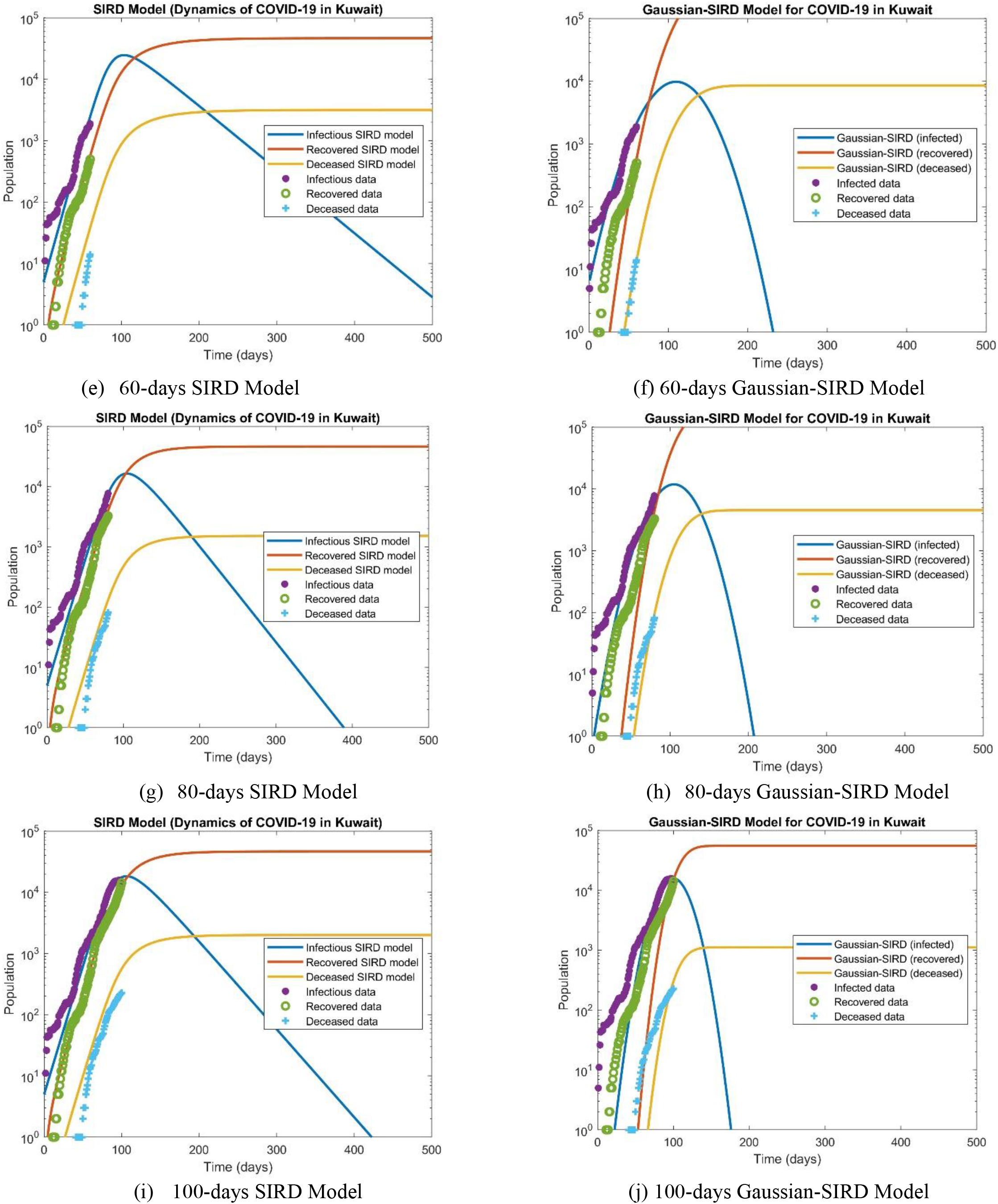
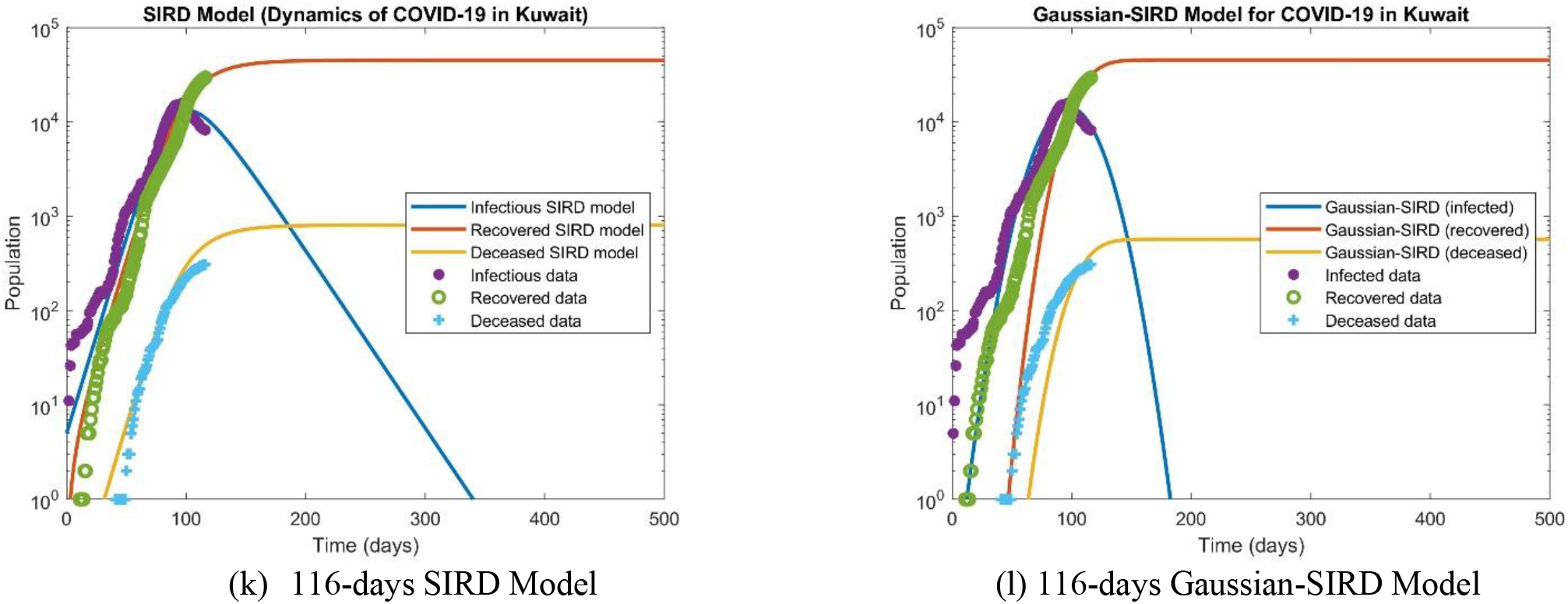
SIRD and Gaussian-SIRD model results for 20, 40, 60, 80, 100, and 116 subsequent days using optimized coefficients for fitting COVID-19 in Kuwait (18 June 2020).

As seen in Figs.1a-1d, the peak day of infection is overpredicted by Gaussian-SIRD model. Gaussian-SIRD model cannot predicts correctly the peak of infection day until 60 days after outbreak. Figs. 1a-1h indicate that SIRD model have started from an overpredicted size of active infected cases until it merges to smaller size; whilst Gaussian-SIRD behaviour is opposite started from small population size to more realistic size after 80 days.

Figs. 1a-1j also indicate that SIRD model have predicted a nearly fixed population size for recovered cases; but Gaussian-SIRD model overpredicted the recovered cases until 100 days from the outbreak. Figs. 1k-1l, shows that both methods have nearly predicated same results after 116 days from outbreak; although better fit is observed using Gaussian-SIRD model to COVID-19 data.

Table 1 shows accuracy and optimized coefficients of SIRD and Gaussian-SIRD equations for 20, 40, 60, 80, 100, and 116 consequent days of COVID-19 data in Kuwait. COVID-19 data were obtained from the ministry of health (MOH) web-link in Kuwait.

**TABLE 1.** Optimized parameters and regression coefficients obtained for SIRD and Gaussian-SIRD models using 20, 40, 60, 80, 100, and 116 subsequent days of COVID-19 in Kuwait (18 June 2020).

| SIRD Model (total population of N=50,000) |  |  |  |  |  |  |  |  |  |  |  |
| --- | --- | --- | --- | --- | --- | --- | --- | --- | --- | --- | --- |
| No. of days | Growth rate ($\beta$) | Recovery rate ($\gamma_R$) | Death rate ($\gamma_D$) | Removing rate ($\gamma$) | Reproduction number ($R_0$) | $R^2(S)$ | $R^2(I)$ | $R^2(R)$ | $R^2(D)$ | | |
| 20 | 0.18311 | 0.01363 | 0.00435 | 0.01798 | 10.18 | - | - | - | - |  |  |
| 40 | 0.14507 | 0.03524 | 0.00215 | 0.03739 | 3.88 | 0.81409 | 0.63761 | 0.95812 | - |  |  |
| 60 | 0.12943 | 0.02308 | 0.00156 | 0.02464 | 5.25 | 0.96106 | 0.93959 | 0.98122 | - |  |  |
| 80 | 0.13963 | 0.04158 | 0.00137 | 0.04295 | 3.25 | 0.99150 | 0.97154 | 0.98236 | 0.57438 |  |  |
| 100 | 0.13399 | 0.03576 | 0.00154 | 0.0373 | 3.59 | 0.99701 | 0.98373 | 0.98172 | 0.70984 |  |  |
| 116 | 0.15188 | 0.05501 | 0.00099 | 0.056 | 2.71 | 0.99870 | 0.91169 | 0.96943 | 0.78406 |  |  |
| Gaussian-SIRD Model (total population of N=700,000) |  |  |  |  |  |  |  |  |  |  |  |
| No. of days | Standard deviation ($\sigma$) | Mean ($\mu$) | Growth rate ($\beta$) | Recovery rate ($\gamma_R$) | Death rate ($\gamma_D$) | Removing rate ($\gamma$) | Reproduction number ($R_0$) | $R^2(S)$ | $R^2(I)$ | $R^2(R)$ | $R^2(D)$ |
| 20 | 48.079 | 157.295 | 0.0608 | 0.8134 | 0.0000 | 0.8134 | 0.0747 | 0.8498 | 0.8008 | 0.7914 | NaN |
| 40 | 49.485 | 161.129 | 0.0685 | 1.412 | 0.0000 | 0.03739 | 0.0485 | 0.9760 | 0.9403 | 0.9596 | NaN |
| 60 | 28.481 | 109.798 | 0.0916 | 0.2532 | 0.0061 | 0.2593 | 0.3533 | 0.9893 | 0.9755 | 0.8920 | 0.9828 |
| 80 | 23.5967 | 105.011 | 0.1068 | 0.1484 | 0.0032 | 0.1516 | 0.7045 | 0.9899 | 0.9627 | 0.8409 | 0.8954 |
| 100 | 17.4553 | 99.1729 | 0.0919 | 0.0396 | 0.0008 | 0.0404 | 2.2748 | 0.9696 | 0.9858 | 0.8957 | 0.7965 |
| 116 | 19.4619 | 97.3516 | 0.0727 | 0.0323 | 0.0004 | 0.0327 | 2.2232 | 0.9899 | 0.9528 | 0.9903 | 0.8654 |

From Table 1, it is observed that for 20 days data SIRD model cannot provide any meaningful value for regression coefficient; however, Gausian-SIRD model cannot provide only for death cases (zero case data). Also, in SIRD model, the total population of N=50,000 is used; but Gaussian-SIRD model requires larger total population size of N=700,000 to properly fit COVID-19 data.

As seen in Table 1, SIRD model gives a large re-production number at start of the pandemic; therefore, it is a best candidate for detecting severity of a pandemic. Gaussian-SIRD method is somehow insensitive in terms of detecting severity of the pandemic and cannot be a good indicator. It gives the re-production number greater than one after 100 days of outbreak. Figure 2 compares results of SIRD and Gaussian-SIRD models on predicting peak day of infection of COVID-19 in Kuwait.

**Figure 2.**
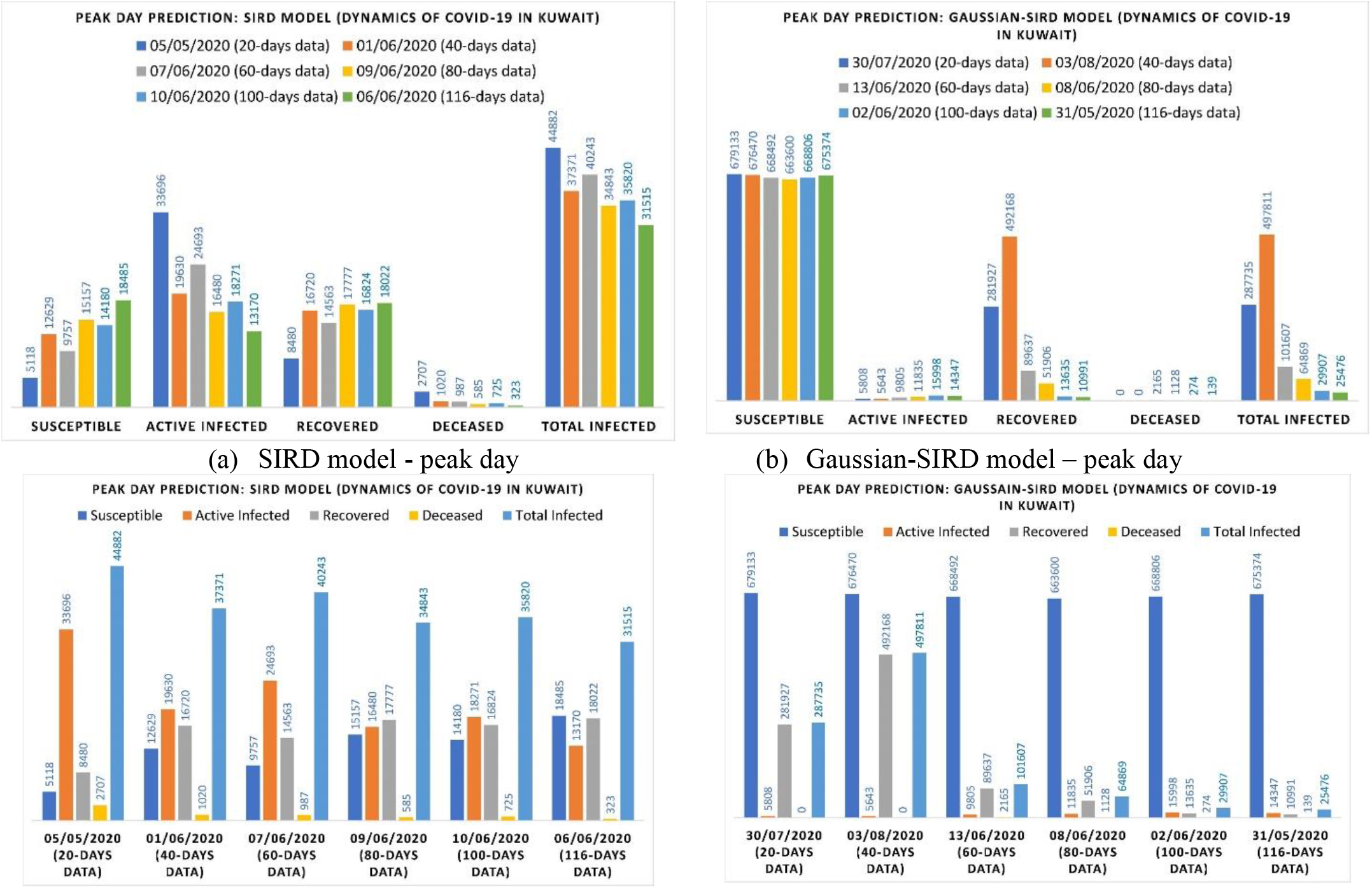
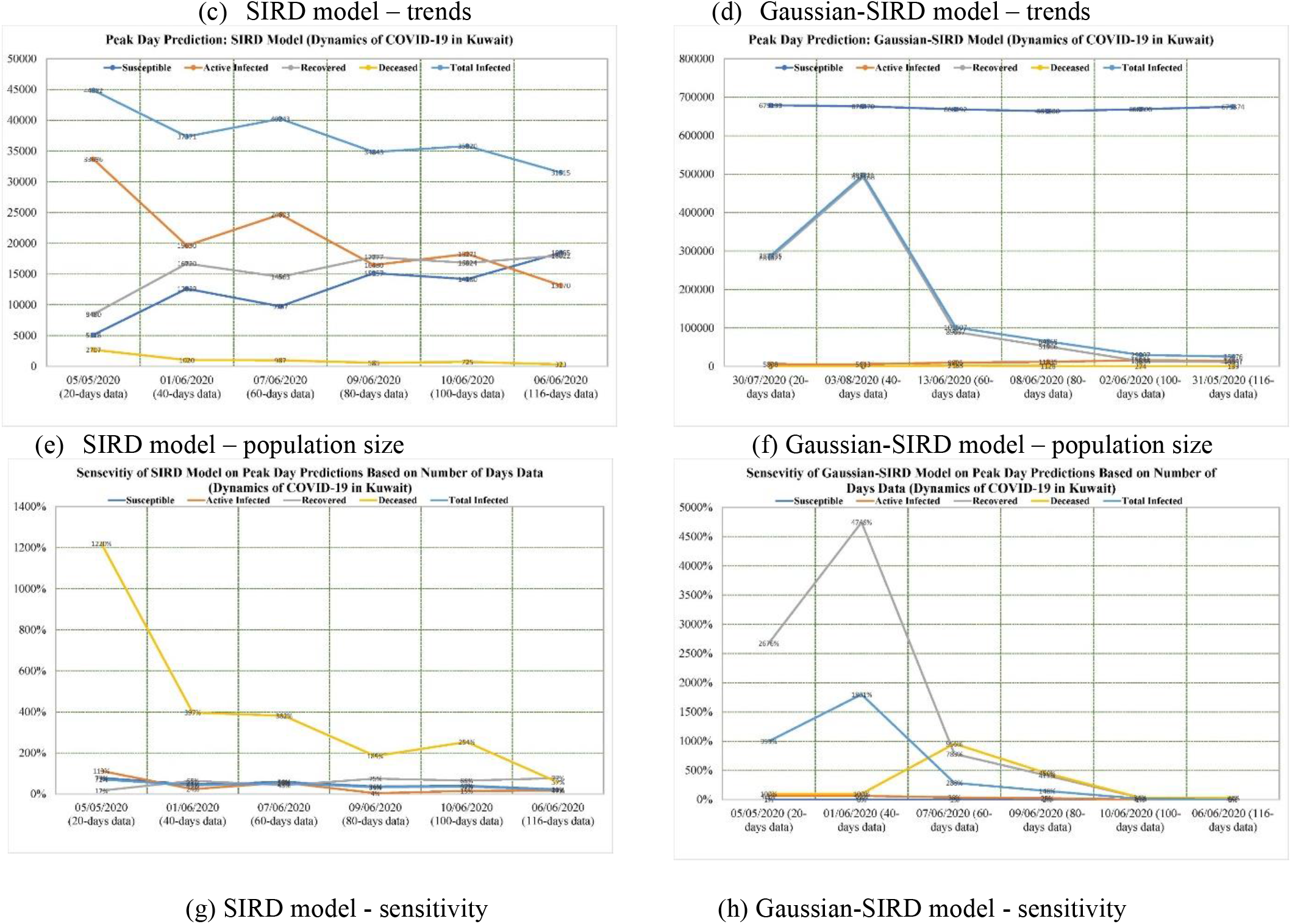
Comparison of SIRD and Gaussian-SIRD models on precision and sensitivity for predicting peak day of infection using 20, 40, 60, 80, 100, and 116 subsequent days of COVID-19 data in Kuwait (18 June 2020).

Fig. 2a shows that SIRD model started with low susceptible, high active infected, low recovered, high deceased and high total infected cases using 20 days of COVID-19 data. The values are gradually moderated as days of pandemic passed by. All high values are giving true warning on spread of disease; although not realistic. In contrast, Gaussian-SIRD model in Fig. 2b started with least alarming results except with large number of total infected cases with zero death cases on peak day of infection. Fig. 2c shows that how SIRD model predictions are moderated gradually by adding more COVID-19 data, whilst Fig. 2d shows that Gaussian-SIRD model have picked up true dynamics of the pandemic after 60 days. Fig. 2e indicates that predicted SIRD populations are widely changes as time evolved; however, Fig. 2f indicate that Gaussian-SIRD model provide more precise prediction on population size after 60 days. Fig. 2g indicates that percentage error of SIRD model remain high for deceased population except after 116 days of data; although other population size prediction errors are not converging to zero after 116 days of data. This is maybe due to high sensitivity of SIRD model to data values than Gaussian-SIRD model (see Fig. 2h). The Gaussian-SIRD model converged to error values below 10% except for deceased population after 116 days. Table 2 compares percentage error on prediction of population sizes on peak day of infection using both methods. Gaussian-SIRD model offers more precise fit after 116 days of COVID-19; yet too late for any practical use.

**TABLE 2.** Sensitivity of SIRD and Gaussian-SIRD models on predicting precise population sizes on peak day of infection as COVID-19 evolved on 20, 40, 60, 80, 100, and 116 subsequent days of the pandemic in Kuwait (18 June 2020).

| Sensitivity of SIRD Model (total population of N=50,000) |  |  |  |  |  |  |  |  |  |  |
| --- | --- | --- | --- | --- | --- | --- | --- | --- | --- | --- |
| No. of days | Susceptible population (S) | Active Infected population (I) | Recovered population (R) | Deceased Population (D) | Total infected cases (I<sub>c</sub>) | Error (%) S | Error (%) I | Error (%) R | Error (%) D | Error (%) I<sub>c</sub> |
| 20 | 5118 | 33696 | 8480 | 2707 | 44882 | 79 | 113 | 17 | 1220 | 71 |
| 40 | 12629 | 19630 | 16720 | 1020 | 37371 | 47 | 24 | 65 | 397 | 43 |
| 60 | 9757 | 24693 | 14563 | 987 | 40243 | 59 | 56 | 43 | 382 | 54 |
| 80 | 15157 | 16480 | 17777 | 585 | 34843 | 36 | 4 | 75 | 185 | 33 |
| 100 | 14180 | 18271 | 16824 | 725 | 35820 | 40 | 15 | 66 | 254 | 37 |
| 116 | 18485 | 13170 | 18022 | 323 | 31515 | 22 | 17 | 77 | 57 | 20 |
| *96 | 23808 | 15831 | 10156 | 205 | 26192 | 0 | 0 | 0 | 0 | 0 |
| Sensitivity of Gaussian-SIRD Model (total population of N=700,000) |  |  |  |  |  |  |  |  |  |  |
| No. of days | Susceptible population (S) | Active Infected population (I) | Recovered population (R) | Deceased Population (D) | Total infected cases (I<sub>c</sub>) | Error (%) S | Error (%) I | Error (%) R | Error (%) D | Error (%) I<sub>c</sub> |
| 20 | 679133 | 5808 | 281927 | 0 | 287735 | 1 | 63 | 2676 | 100 | 999 |
| 40 | 676470 | 5643 | 492168 | 0 | 497811 | 0 | 64 | 4746 | 100 | 1801 |
| 60 | 668492 | 9805 | 89637 | 2165 | 101607 | 1 | 38 | 783 | 956 | 288 |
| 80 | 663600 | 11835 | 51906 | 1128 | 64869 | 2 | 25 | 411 | 450 | 148 |
| 100 | 668806 | 15998 | 13635 | 274 | 29907 | 1 | 1 | 34 | 34 | 14 |
| 116 | 675374 | 14347 | 10991 | 139 | 25476 | 0 | 9 | 8 | 32 | 3 |
| *96 | 673808 | 15831 | 10156 | 205 | 26192 | 0 | 0 | 0 | 0 | 0 |

\* Actual data on peak day

## IV. Conclutions

Construction of analytical solutions to SIR type models may provide quick and precise guide on development of endemic/pandemic diseases to assist health organization and policy makers on controlling crisis. In this paper, a new method is introduced to integrate Gaussian distribution function within SIRD model to study endemic/pandemic data. Gaussian distribution function is a widely used statistical tool in natural and social sciences. Sensitivity and precision of both SIRD and Gaussian-SIRD methods are investigated here using COVID-19 data in Kuwait and the following conclusions are drawn:

- Accuracy trends of SIRD model using 20, 40, 60, 80, 100, and 116 days of COVOD-19 data in Kuwait are assessed and it is found that sensitivity of SIRD model is high which gives quickly right alarm even with 20 days of data.
- Gaussian-SIRD model requires a large susceptible population to accurately track development of COVID-19; yet the sensitivity of model is very low to be considered as a quick tool for detecting alarming condition.
- SIRD model predictions on size of exposed population is not very accurate and error percentage remain high even after 116 days of COVID-19 outbreaks.
- Gaussian-SIRD model showed some merits in terms of precision provided below 10% error on population sizes after 116 days of COVID-19 outbreak.

Integrating suitable probability density functions within SIR type models provide analytical solutions to epidemic/pandemic situations. These analytical solutions can be easily optimized to fit epidemic/pandemic data to provide fast and robust predictions..

## Data Availability

Data Online available here:
https://www.worldometers.info/coronavirus/

https://www.worldometers.info

